# Cohort profile: The Nanjing Diabetes Cohort database – a population-based surveillance cohort

**DOI:** 10.1101/2024.02.05.24302328

**Authors:** Wei Feng, Yuechuchu Yin, Chunyan Gu, Yuan Mu, Duxiao Zhang, Zhenhuan Tao, Weidong Yin, Xin Zhang, Yun Yu, Jie Hu, Cheng Wan, Yun Liu

## Abstract

**Purpose:** To study epidemiology, complications, risk factors, clinical course and treatment patterns of diabetes, the Nanjing Diabetes Cohort (NDC) was established using anonymized electronic medical records from 650 hospitals and primary cares since 2020. This cohort provides valuable data for researchers and policymakers working on diabetes management and public health strategies.

**Participants:** Diabetes was defined as having inpatient or outpatient encounters with diagnosis of diabetes ICD-9/10 codes, any use of insulin or oral hypoglycemic drugs, or one encounter with hemoglobin A1C > 4.8mmol/mol or 6.5%. Patients with diabetes have been continuously enrolled from hospitals and primary cares in Nanjing since 2020. Demographic, medications, and comorbidities data were extracted using clinical notes, diagnostic codes, labs, medications, and vital signs among different types of diabetes.

**Findings to date:** The NDC consisted of 1033904 patients from Jan 1^st^, 2020 to Dec 31th, 2022, the most part of whom were male (50.62%) and from district Gulou (30.79%). The clinical characteristics and medications usage of patients with type 1 diabetes, type 2 diabetes, gestational diabetes and other diabetes were assessed. The prevalence of hypertension, heart failure, and cerebrovascular disease were 31.20%, 21.46%, and 13.74% respectively.

**Future plans:** NDC will enroll eligible patients continuously each year. The data of NDC is maintained by Department of Medical Informatics, Nanjing Medical University.

**Strengths and limitations of this study:**

1. The NDC is an ongoing population-base, large-sized cohort that contains over 1000000 patients with type 1 diabetes, type 2 diabetes, gestational diabetes, and other diabetes with follow-up records.
2. NDC covers the patients with diabetes from Nanjing with urban and suburban areas, guaranteeing the generalizability of coverage.
3. The NDC captures people with diabetes continuously from the Nanjing healthcare information platform, and provides exhaustive clinical data on clinical report, laboratory results, glucose-lowering drugs, diabetes-related comorbidities, and death report.
4. Patients who had done no measurement of glycemic indexes or suffered prediabetes with normal HbAlc would not be included in the NDC.

**Data availability statement:** Data are available on reasonable request. Data may be made available from the corresponding author with reasonable request, pending institutional and IRB approval and after completing a data sharing agreement.

**Funding declaration:** This study was supported by the industry prospecting and common key technology key projects of Jiangsu Province Science and Technology Department (Grant no. BE2020721), the industrial and information industry transformation and upgrading special fund of Jiangsu Province in 2021 (Grant no. [2021]92), the key project of smart Jiangsu in 2020 (Grant no. [2021]1), Jiangsu Province Engineering Research Center of big data application in chronic disease and intelligent health service (Grant no. [2020]1460).

**Collaborators:** The data of the NDC are not open access but can be shared under conditions of collaboration and endowment. Collaborative research projects are encouraged. For more detailed information on the NDC, please contact: Prof. Yun Liu

**Contributors:** Study concept and design: Wei Feng, Yuechuchu Yin, Chen Wan; acquisition of data: Wei Feng, Yuechuchu Yin, Chunyan Gu, Chen Wan, Yuan Mu, Zhenhuan Tao, Weidong Yin; data analysis and verification: Chunyan Gu, Xin Zhang, Yuzhuo Wang, Yun Yu, Yuan Mu, Duxiao Zhang; drafting of the manuscript: Wei Feng, Yuechuchu Yin, Chen Wan; acquisition of funding: Yun Liu. All authors have read and approved the final version.

**Competing interests:** None declared.

**Patient consent for publication:** Not required.

**Ethics approval:** The study was approved by the ethical review board of the First Affiliated Hospital, Nanjing Medical University (IRB#2019-SR-153).

## Introduction

Diabetes can cause a series of complications and comorbidities including high blood pressure, depression, stroke, heart attack, retinopathy, and infections[1,2]. The population-based cohorts from real-world settings can help to study epidemiology, complications, risk factors, clinical course and treatment patterns of diabetes[3]. Moreover, while most people with diabetes are in low-income and middle-income settings, most population-based studies and data resources are in high-income settings[4].

Large diabetes databases can help researchers improving the knowledge on strategies of diabetes prevention and diabetes epidemiological research. In China, the number of people with diabetes is 116.4 million and will reach 140.5 million in 2030[5]. According to a national cross-sectional study, the diabetes prevalence was increased from 10.9% (2013) to 12.4% (2018) in China[6]. There are several large population-based cohorts launched in China. China Health and Nutrition Survey (CHNS) is a nationally ongoing open cohort that was initialed in 1989[7]. It provides information of numerous changes in socioeconomic areas of families throughout 12 provinces in China. Risk Evaluation of Cancers in Chinese Diabetic Individuals, a Longitudinal (REACTION) study is aimed to explore the association of type 2 diabetes and pre-diabetes with the risk of cancer since 2011[8]. The Hong Kong Diabetes Surveillance Database (HKDSD) focus on the epidemiology of gestational diabetes, prediabetes and diabetes in Hong Kong, China[9]. It captured clinical information from electronic medical system since 2000.

Nanjing is the capital of Jiangsu Province, with 9.31 million residents according to data from China’s seventh national census in 2021[10]. Nanjing is one of the most comprehensive industrial production bases and an important transportation hub in East China[11]. From 2011 to 2013, a survey on total of 4918 adult residents in Nanjing showed that the prevalence rate of metabolic syndrome was 26.3%[12]. The prevalence rates in 2 districts of Nanjing were estimated to be 6.0% for diabetes in 2017[13], which was lower than the national average (11.2%)[14]. Therefore, it is crucial to identify the diabetes risks and improve knowledge on strategies and epidemiological status for diabetes in Nanjing using large population-based cohorts.

The Nanjing Diabetes Cohort (NDC) is an electronic record of medical encounters, including all patients who meet the diabetes criteria since 2020 in health services providers of Nanjing. The NDC was established with the major aims of: (1) integrating EHR data for diabetes and complications in Nanjing; (2) exploring the clinical factors on diabetes progression; (3) developing individualized intervention strategies for the treatment of diabetes and comorbidities.

## Cohort Description

### Study setting and participants

In 2017, the Nanjing healthcare information platform was started to routinely collect clinical information for all people attending hospitals or primary healthcare institutions. As of 2020, there were 650 medical institutions from 11 districts in NDC (Figure 1). A unique identification number is used in the platform to link EHR and other data sets, such as mortality records. To analyze the data anonymously, a unique number generated by platform is assigned to each patient.

**Figure 1.**
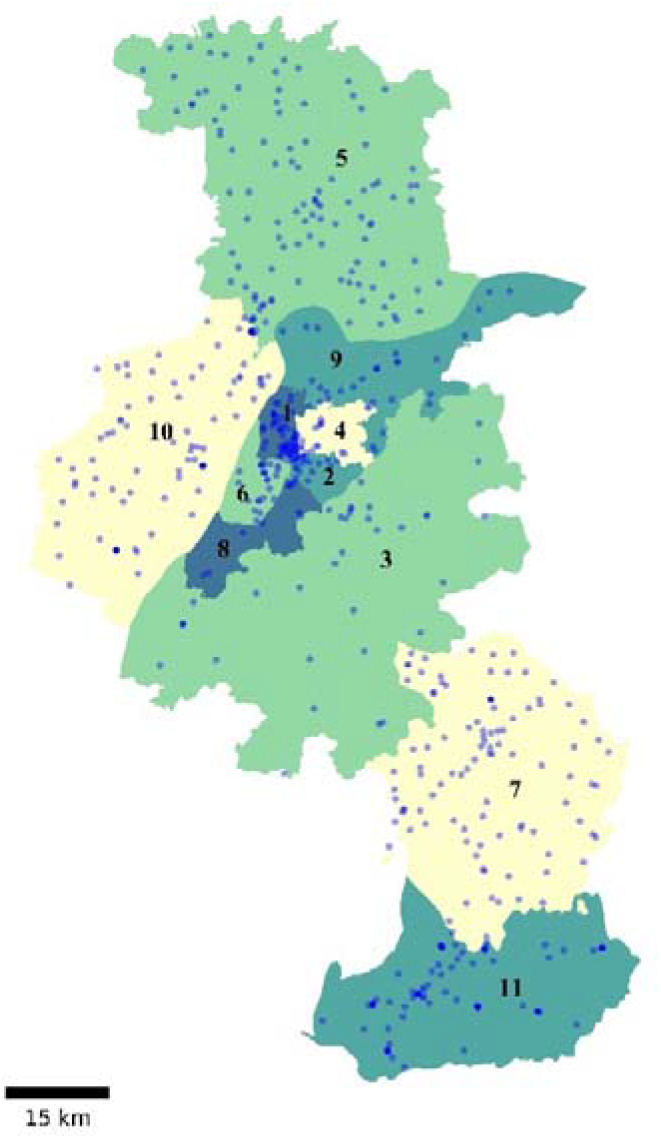
Distribution of medical institutions in NDC from 11 districts of Nanjing. Districts (sorted by the number of patients in each district): 1, Gulou; 2, Qinhuai; 3, Jiangning; 4, Xuanwu; 5, Luhe; 6, Jianye; 7, Lishui; 8, Yuhuatai; 9, Qixia; 10, Pukou; 11, Gaochun.

We defined diabetes in anyone with any of the following three criteria (1) at least one encounter with haemoglobin A1c (HbA1c) level ≥6.5%[16] or (2) any encounters (inpatient or outpatient) with diagnosis of diabetes or (3) any encounters with prescriptions of diabetes medications. These were based upon the Chinese Diabetes Society (CDS)[15] and the “Standards of Medical Care in Diabetes” by American Diabetes Association (ADA)[16]. According to the CDS and ADA recommendations [15][17], diabetes medications include: dipeptidyl peptidase 4 (DPP-4) inhibitor, glucagon-like peptide 1 (GLP-1) receptor agonist (RA), α-glucosidase inhibitors, insulin, biguanides, sodium–glucose cotransporter 2 (SGLT2) inhibitor, sulphonylureas, thiazolidinedione, and traditional Chinese medicine (TCM).

### Data collection

The dataset includes all encounters on or after 1^st^ January 2020 covering 11 districts (Figure 1). The 3 districts which most data came from were Gulou (30.79%), Qinhuai (21.99%), and Jiangning (11.95%). The cohort is ongoing with continuous enrolment annually.

Each participant has a unique clinical identifier. In Table 1, features for individual-level data in NDC including demographics, clinic visits for outpatients, admissions and discharges for inpatients, diagnoses, laboratory measurements and medications which were obtained from the platform. Demographics including sex and date of birth are static variables. Laboratory measurements, medications, principal and secondary diagnosis codes are captured from both outpatients and inpatients. All patients are followed until death. The cause of death is stored as death report.

**Table 1.** Summary of measurements for patients in NDC.

| Category | Measurements |
| --- | --- |
| Demographics | sex, date of birth |
| Outpatient clinic attendance | principal/secondary diagnosis codes (ICD-10) |
| Hospital admission | admission report, principal/secondary diagnosis codes of admission (ICD-10) |
| Hospital discharge | admission report, discharge report, principal/secondary diagnosis codes of discharge (ICD-10), death report |
| Glycaemic indices | FPG, HbA1c, random and after-meal glucose |
| Lipid profile | TC, LDL, HDL |
| Kidney function | ALB, eGFR, Urine ACR |
| Medications | DPP-4 inhibitor, GLP-1 RA, α-glucosidase inhibitors, insulin, biguanides, SGLT2 inhibitor, sulphonylureas, thiazolidinedione, glinide, and TCM |
| Clinical notes | complaint, history of illness |
| General information for diabetes | classification of diabetes, BMI |

FPG, fasting plasma glucose; TC, total cholesterol; LDL, low-density-lipoprotein cholesterol; HDL, high-density-lipoprotein cholesterol; ALB, urine trace albumin; eGFR, estimated glomerular filtration rate; ACR, ratio of urinary albumin and creatinine; BMI, body mass index; TCM, traditional Chinese medicine.

Laboratory measurements used in clinical encounters from the platform are automatically captured via clinical identifier number. These measurements include glycaemic indices (FPG, HbAlc, random and after-meal glucose), lipid profile (TC, LDL, and HDL), kidney function (ALB, eGFR, and Alb/Cr). Diagnosis codes during the encounter of hospitalizations or clinic visits are coded using ICD-10. Each encounter contains a principal diagnosis code and less than 9 secondary diagnosis code.

The NDC also captures diabetes comorbidities or complications, including hypertension, cerebrovascular disease (CVD), angina pectoris, myocardial infarction, heart failure, mental disorders (depression and anxiety), and other diabetes-related conditions (diabetic foot, diabetic retinopathy, kidney failure, ketoacidosis, and severe hypoglycaemia). For the screening criteria and coding setup of the aforementioned diabetic complications, please refer to the Table S1 in Supplemental File 2.

The medications for each encounter are also captured by NDC. Data are available on the type, dose, and fee of treatment with glucose-lowering drugs, including DPP-4 inhibitor, GLP-1 RA, α-glucosidase inhibitors, insulin, biguanides, SGLT2 inhibitor, sulphonylureas, thiazolidinedione, glinide, and TCM. The names and categories of each drug can be found in Table S2 Supplemental File 2.

The classification of diabetes is processed through diagnosis, present history of diseases, and medication usage from platform. For detailed procedures on diabetes classification, please refer to the Figure S1 in Supplemental File 2. Total of 1033904 participants with diabetes were included from platform after criteria of diabetes applied. Participants are classified into: type 1 diabetes mellitus (T1DM), type 2 diabetes mellitus (T2DM), gestational diabetes mellitus (GDM), and other diabetes, according to classification approach from CDS[15] and ADA[16]. GDM, T1DM, and other diabetes (monogenic diabetes, pancreatic diabetes, steroid diabetes, or treated with glucocorticoid/interferon α) were checked through diagnosis and history of diseases. To identify all patients with T2DM, patients aged >= 30 who diagnosed with diabetes related disease, has diabetes-related history, or prescribed at least one non-insulin medications before insulin were also classified as T2DM. At last, 20341 participants were classified as unclear.

## Findings to date

The characteristics of the cohort recruited between 1^st^ Jan, 2020 and 31th Dec, 2022 are summarized in Table 2. Overall, 94.58% (n=977880) of patients had T2DM, 0.47% (n=4818) had T1DM, 2.16% (n=22331) had GDM, 0.83% (8534) had other diabetes, and 1.97% (n=20341) is unknown due to the lack of diabetes-related information. Notably, 80.03% of patients in unclear group were female, and the mean of FBG was 7.7 (SD: 2.82). Most patients in the “unclear group” could have GDM. The overall description of the data is detailed in Supplemental File 1.

**Table 2.**
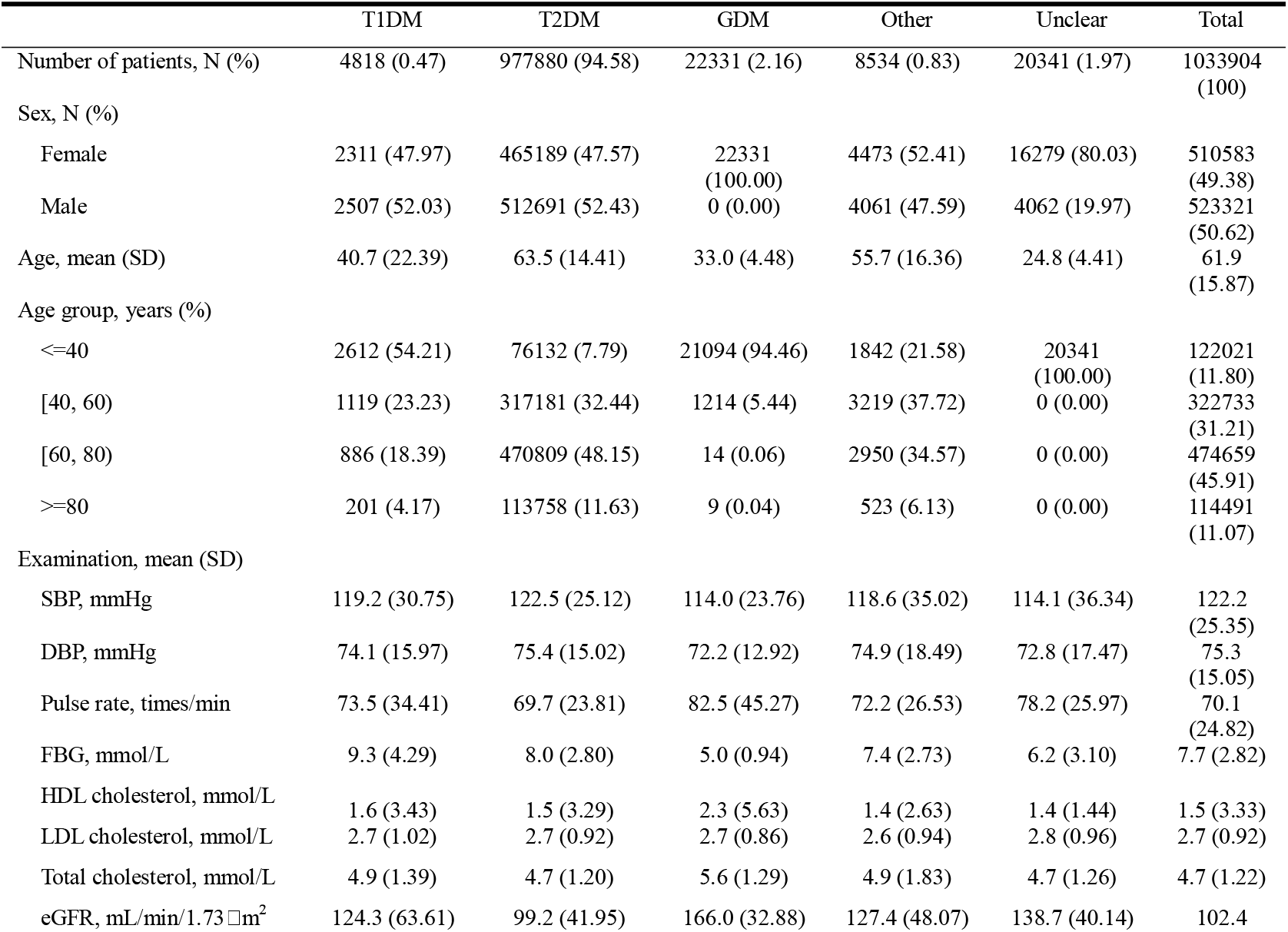
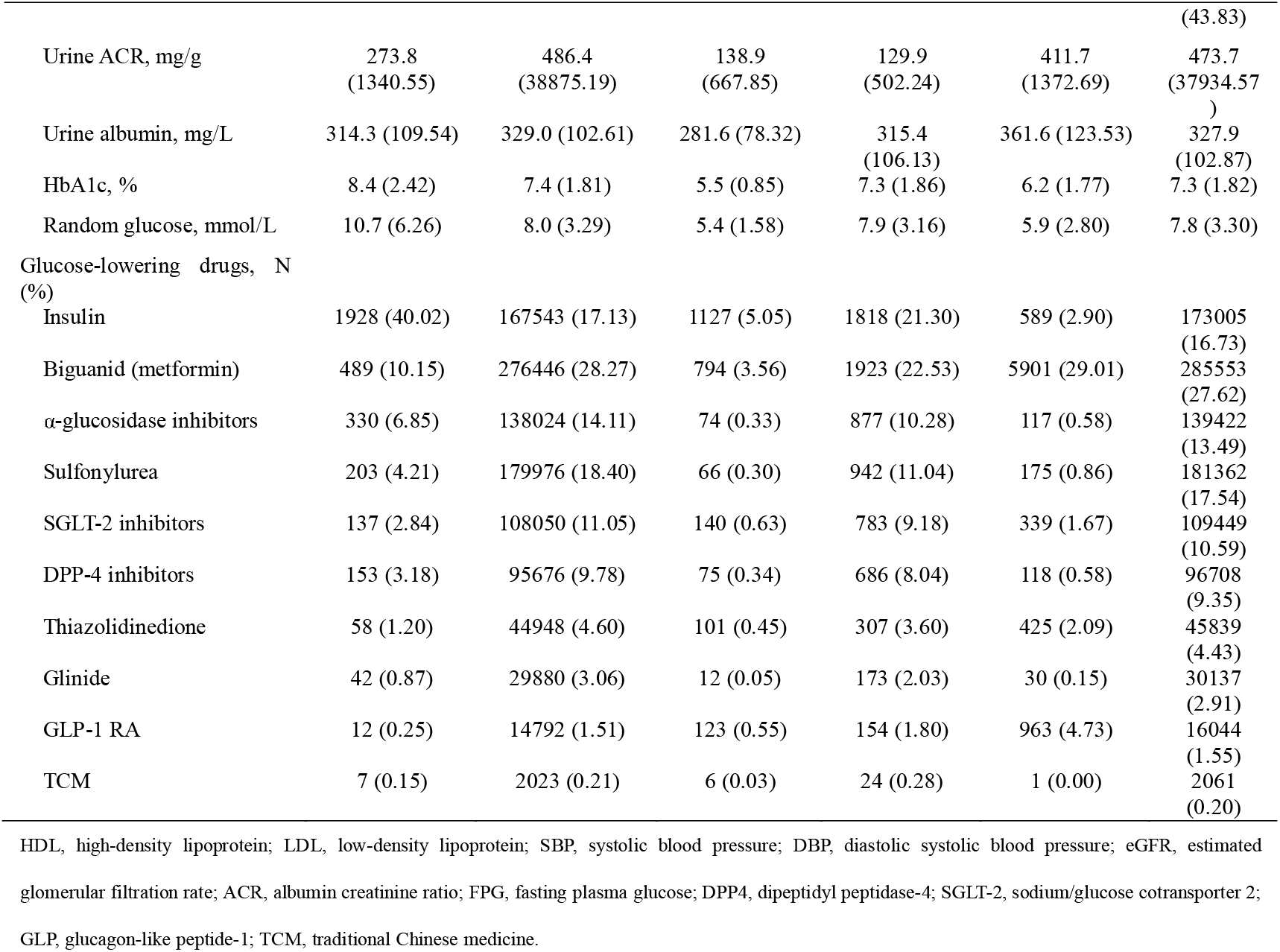
Baseline characteristics of patients according to diabetes type in NDC, 2020-2022.

For patients with T1DM or T2DM, over 50% were male. The percentage of men was higher among patients with T2DM (52.43%) in the cohort than the rest types of diabetes, which is comparable to a Chinese cross-sectional study in 2018[14]. Mean age of patients in GDM (33 years) and T1DM (40 years) group were lower than T2DM (63.5 years). This likely reflects the classification process of patients for T2DM. About 7.79% (n=76132) of T2DM patients were under 40, which is lower than the early onset of diabetes in Beijing (10.9%)[18]. This might result from environmental and socioeconomic of different regions and cities[6,14,19].

Mean DBP levels were similar among patients with T2DM (75.4 mm Hg, SD: 15.02), T1DM (74.1 mm Hg, SD: 15.97), and other diabetes (74.9 mm Hg, SD: 18.49). And SBP showed the similar pattern, while it was much lower in GDM (114.0 mm Hg, SD: 23.76). Among patients with GDM, Mean HDL (2.3 mmol/L, SD: 5.63) and Total cholesterol level (5.6 mmol/L, SD: 1.29) were higher than other types of diabetes. The mean HbA1c were higher in T1DM (8.4%, SD: 2.42) compared to T2DM (7.4%, SD: 1.81). Markedly elevated glucose levels are often showed in patients with T1DM[20]. Also, mean random glucose level was higher in patients with T1DM (10.7 mmol/L, SD: 6.26) than T2DM (8.0 mmol/L, SD: 3.29). In addition to insulin treatment (16.73%), majority of prescriptions were on first-line and second-line medications (metformin and sulfonylurea) in T2DM.

As shown in Table 3, the prevalence rate per 1000 person-years of hypertension is higher among patients with T2DM (297.68) than the other comorbidities, which is comparable to a Chinese cohort[21]. The prevalence rates of angina pectoris (58.43) and CVD (147.46) were higher in T2DM compared to T1DM. While the prevalence rates of diabetic retinopathy and ketoacidosis were higher in T1DM.

**Table 3.**
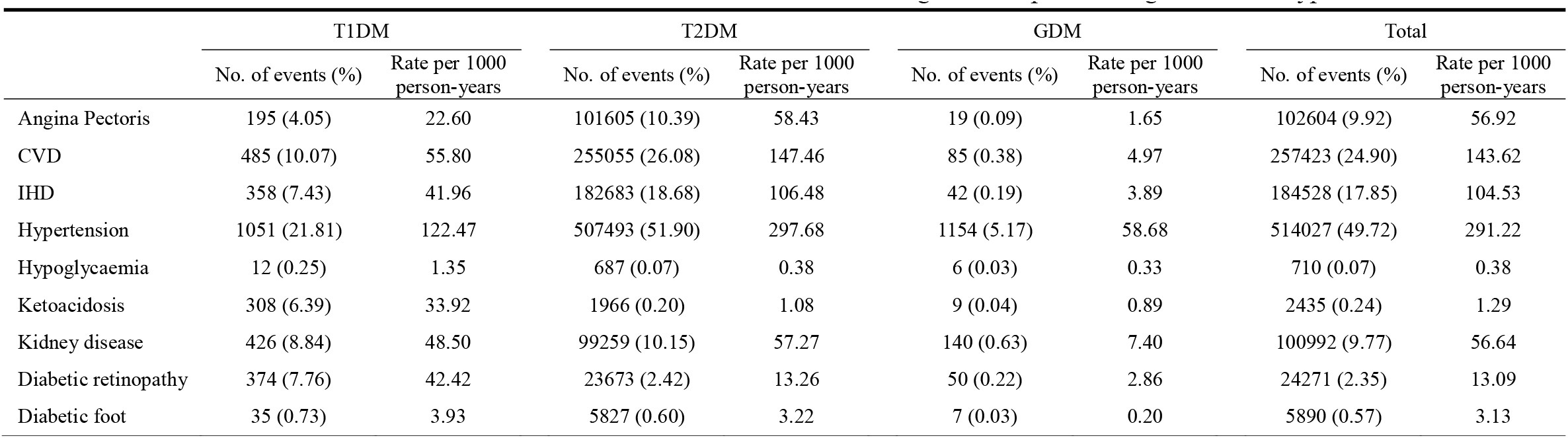
Number of and incidence rates for diabetes-related conditions during follow-up according to diabetes type, 2020-2022.

|  | T1DM |  | T2DM |  | GDM |  | Total |  |
| --- | --- | --- | --- | --- | --- | --- | --- | --- |
|  | No. of events (%) | Rate per 1000 person-years | No. of events (%) | Rate per 1000 person-years | No. of events (%) | Rate per 1000 person-years | No. of events (%) | Rate per 1000 person-years |
| Angina Pectoris | 195 (4.05) | 22.60 | 101605 (10.39) | 58.43 | 19 (0.09) | 1.65 | 102604 (9.92) | 56.92 |
| CVD | 485 (10.07) | 55.80 | 255055 (26.08) | 147.46 | 85 (0.38) | 4.97 | 257423 (24.90) | 143.62 |
| IHD | 358 (7.43) | 41.96 | 182683 (18.68) | 106.48 | 42 (0.19) | 3.89 | 184528 (17.85) | 104.53 |
| Hypertension | 1051 (21.81) | 122.47 | 507493 (51.90) | 297.68 | 1154 (5.17) | 58.68 | 514027 (49.72) | 291.22 |
| Hypoglycaemia | 12 (0.25) | 1.35 | 687 (0.07) | 0.38 | 6 (0.03) | 0.33 | 710 (0.07) | 0.38 |
| Ketoacidosis | 308 (6.39) | 33.92 | 1966 (0.20) | 1.08 | 9 (0.04) | 0.89 | 2435 (0.24) | 1.29 |
| Kidney disease | 426 (8.84) | 48.50 | 99259 (10.15) | 57.27 | 140 (0.63) | 7.40 | 100992 (9.77) | 56.64 |
| Diabetic retinopathy | 374 (7.76) | 42.42 | 23673 (2.42) | 13.26 | 50 (0.22) | 2.86 | 24271 (2.35) | 13.09 |
| Diabetic foot | 35 (0.73) | 3.93 | 5827 (0.60) | 3.22 | 7 (0.03) | 0.20 | 5890 (0.57) | 3.13 |

Table 4 show the prevalence of patients in NDC from years 2020 to 2022. The number of diabetic patients in NDC had kept growing in the past 3 years. Among them, the number of T2DM patients increased from 529265 in 2020 to 660968. However, due to the uncertainty about the types of diabetes from some patients, we assumed patients over 30 years had T2DM.

**Table 4.** Data characteristics of patients in the NDC dataset, 2020-2022.

|  | 2020 | 2021 | 2022 |
| --- | --- | --- | --- |
| Patients, N | 544421 | 638319 | 689340 |
| Gender, N (%) |  |  |  |
| Female | 260759 (47.90) | 311475 (48.80) | 332506 (48.24) |
| Male | 283662 (52.10) | 326844 (51.20) | 356834 (51.76) |
| Age, mean (SD) | 64.3 (14.50) | 63.5 (14.80) | 63.7 (14.96) |
| Age Group, N (%) |  |  |  |
| (0, 40] | 39484 (7.25) | 55476 (8.69) | 59344 (8.61) |
| (40, 60] | 160746 (29.53) | 189227 (29.64) | 203771 (29.56) |
| (60, 80] | 276838 (50.85) | 321656 (50.39) | 345126 (50.07) |
| (80, 1000] | 67353 (12.37) | 71960 (11.27) | 81099 (11.76) |
| Diabetes Type, N (%) |  |  |  |
| T1DM | 2693 (0.49) | 3055 (0.48) | 3266 (0.47) |
| T2DM | 529265 (97.22) | 610005 (95.56) | 660968 (95.88) |
| GDM | 1768 (0.32) | 12549 (1.97) | 11069 (1.61) |
| Other | 4413 (0.81) | 5156 (0.81) | 5398 (0.78) |
| Unknown | 6282 (1.15) | 7554 (1.18) | 8639 (1.25) |
| Laboratory records, mean (SD) |  |  |  |
| SBP, mmHg | 121.6 (26.65) | 123.2 (24.85) | 122.9 (24.00) |
| DBP, mmHg | 75.0 (15.95) | 75.7 (14.52) | 75.2 (14.27) |
| Pulse rate, times/min | 68.4 (21.18) | 69.0 (21.46) | 71.8 (25.00) |
| FBG, mmol/L | 8.0 (2.72) | 7.6 (2.67) | 7.7 (2.74) |
| HDL cholesterol, mmol/L | 1.8 (4.32) | 1.5 (3.14) | 1.4 (2.86) |
| LDL cholesterol, mmol/L | 2.7 (0.92) | 2.7 (0.93) | 2.6 (0.92) |
| Total cholesterol, mmol/L | 4.6 (1.19) | 4.7 (1.24) | 4.7 (1.24) |
| eGFR, mL/min/1.73 m<sup>2</sup> | 112.2 (41.50) | 101.0 (44.88) | 97.5 (43.98) |
| Urine ACR, mg/g | 765.4 (64999.12) | 348.7 (1157.97) | 315.8 (1108.40) |
| Urine albumin, mg/L | 329.7 (103.86) | 327.0 (102.59) | 330.2 (101.54) |
| HbA1c, % | 7.2 (1.77) | 7.3 (1.86) | 7.4 (1.79) |
| Random glucose, mmol/L | 7.6 (3.37) | 7.9 (3.30) | 8.1 (3.31) |
| Diabetes-related conditions , N (%) |  |  |  |
| Angina Pectoris | 46781 (8.59) | 55690 (8.72) | 43442 (6.30) |
| CVD | 124905 (22.94) | 142007 (22.25) | 105000 (15.23) |
| Heart Failure | 3754 (0.69) | 5055 (0.79) | 3472 (0.50) |
| IHD | 90743 (16.67) | 124808 (19.55) | 89872 (13.04) |
| Hypertension | 289760 (53.22) | 322623 (50.54) | 246383 (35.74) |
| Hypoglycaemia | 223 (0.04) | 275 (0.04) | 213 (0.03) |
| Ketoacidosis | 635 (0.12) | 876 (0.14) | 588 (0.09) |
| Kidney disease | 43306 (7.95) | 68175 (10.68) | 52214 (7.57) |
| Diabetic retinopathy | 7763 (1.43) | 11253 (1.76) | 8332 (1.21) |
| Diabetic foot | 1498 (0.28) | 2563 (0.40) | 1709 (0.25) |

## Strengths and limitations

There are several strengths of the NDC cohort. First, the NDC is an ongoing population-base, large-sized cohort that contains over 550000 patients with type 1 diabetes, type 2 diabetes, gestational diabetes, and other diabetes with follow-up records. It would provide data for future studies of diabetes duration with medication evaluation for a large population. Second, the NDC coverts the diabetes patients from Nanjing with urban and suburban areas, guaranteeing the generalizability of coverage. Each diabetic patient has longitudinal reliable medical records, including medical record data that had been verified manually. Third, the NDC captures patients with diabetes continuously from the Nanjing healthcare information platform, and provides exhaustive clinical data on clinical report, laboratory results, glucose-lowering drugs, diabetes-related comorbidities, and death report.

The NDC also has several limitations. First, patients who had done no measurement of glycemic indexes or suffered prediabetes with normal HbAlc would not be included in the NDC. Second, given the nature of administrative data, not everyone can complete all measurements. The missing variables are inevitable in EHR. Third, NDC captured data from health services providers in Nanjing only. Patients who have emigrated out of Nanjing could censor the follow-up.

## Follow-up schedule

The cohort follow-ups will consist of annual checks of Nanjing healthcare information platform. The baseline data was linked to the platform. All encounters from the same participants and new participants who meet the criteria of diabetes would be captured. In order to keep track patients’ medical events and medication usage, the NDC will censor only at death.

## Patient and public involvement

No patient involved.

## Acknowledgements

We thank all NDC participants, the former and present researchers in this study. We also thank all the members severed in the Nanjing Health Information Center.

