## Supplemental File 2 for "Cohort profile: The Nanjing Diabetes Cohort database – a population-based surveillance cohort"

The classification process for patients with diabetes is depicted in the following diagram (Figure S1). This procedure is carried out annually on the data platform of the Nanjing Health Information Center, where data on diabetic patients is selected based on their medical records and other pertinent information, to conduct a precise analysis of the type of diabetes. The classification process remains consistent each year as delineated in the green-framed flowchart below: initially, all subjects are evaluated for Gestational Diabetes Mellitus (GDM), after which records with incorrect gender data are excluded. Subsequently, a determination is made for other types of diabetes, such as pancreatic diabetes and other endocrine diseases. The subsequent step involves assessing the diagnosis of Type 1 Diabetes Mellitus (T1DM). Among the remaining patients, it is then considered whether a diagnosis of Type 2 Diabetes Mellitus (T2DM) exists. The patients who remain unclassified and are over the age of 30 with a diagnosis of diabetic-related comorbidities or those on non-insulin antidiabetic drugs are categorized as T2DM; the rest are marked as Unclear.

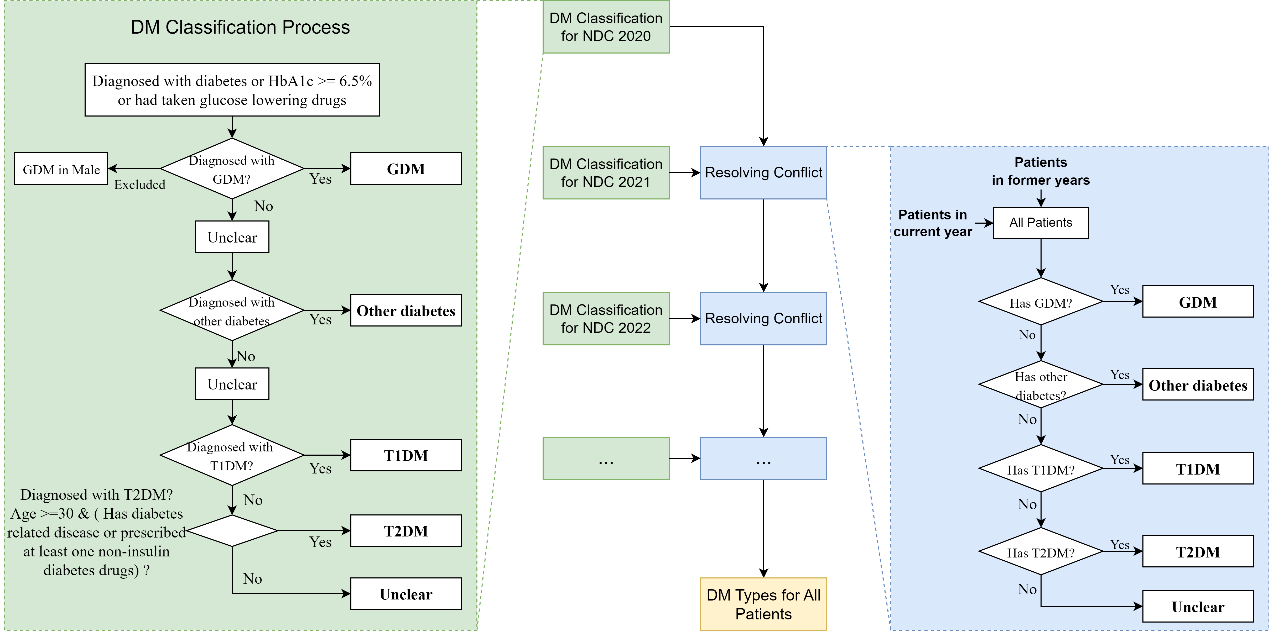

Figure S1 Classification process of diabetes type

Considering that some patients may be diagnosed with a different type of diabetes in the subsequent year, after completion of the annual data classification, a conflict resolution process is undertaken as shown on the right side of the diagram. This process involves merging data of patients previously marked with a type of diabetes from the previous year with the current patient database. A priority-based determination is then conducted for the presence of GDM, Other diabetes, T1DM, and T2DM labels, with the remainder marked as Unclear.

By repetitively cycling through the aforementioned two-step process each year, we aim to generate a comprehensive and meticulously classified diabetes patient type database.

Table S1 Disease screening and coding criteria

| Diseases | Keywords/Regex Expression in Chinese | ICD-10 Codes |
| --- | --- | --- |
| Depression | 抑郁，忧郁，双相 | F31-F34，F39，F06.3 |
| Anxiety | 焦虑 | F40-F43，F06.4 |
| Hypertension | 高血压 | I10-I13，I15 |
| Myocardial Infarction | 心肌缺血，缺血性心，心肌梗，心绞痛 | I20-I25 |
| Heart Failure | 心功能，心衰，心力衰 |  |
| Diabetic Foot | 糖尿病性?足 |  |
| Diabetic Retinopathy | 糖尿病[^#]*?视网膜 |  |
| Parkinson Disease | 痴呆，帕金森病，阿尔茨海默，阿耳茨海默 | F00-F03，G30，F05.1，G31.1 |
| Cerebrovascular Disease | 中风，脑卒中，((脑(半球)?)\|(腔(隙性?)?)\|(颈内?)\|(枕叶)\|(额叶)\|(顶叶)\|(颅内?)\|(脑膜)\|(硬膜)\|(蛛网膜?)\|(基底(节区)?)\|(颞叶)).?((多发性?)\|(陈旧性?)\|(循环)\|([0-9a-zA-Z\-]+(段\|(交界区?))))?(((出\|缺)血)\|(梗)\|(血肿)\|(栓塞)\|(狭窄)\|(动脉)\|(血管)\|(血栓)\|(静脉))  ((硬化)\|(缺血)\|(缺氧))性?脑病 | G45，G46，I60-I69，H34.0，E14.5 |
| Kidney Disease | 糖尿病[^#]*?肾，慢性肾 | N03-N05，N18，N19，N25，I12，I13，E14.2 |

Table S2 Antidiabetic Drugs

| Antidiabetic Drugs | Drug Names |
| --- | --- |
| DPP-4 inhibitor | Saxagliptin, Linagliptin, Alogliptin, Sitagliptin, Vildagliptin. |
| GLP-1 RA | Dulaglutide, Lixisenatide, Liraglutide, Exenatide. |
| α-Glucosidase inhibitors | Acarbose, Voglibose, Miglitol. |
| Insulin | Insulin. |
| Biguanides | Metformin. |
| SGLT2 inhibitor | Dapagliflozin, Empagliflozin, Canagliflozin. |
| Sulphonylureas | Glimepiride, Glipizide, Gliclazide, Glyburide, Glipentide. |
| Thiazolidinedion | Rosiglitazone, Pioglitazone. |
| Glinide | Nateglinide, Repaglinide, Mitiglinide. |
| TCM | Xiaoke |
